# Nirmatrelvir plus ritonavir for early COVID-19 and hospitalization in a large US health system

**DOI:** 10.1101/2022.06.14.22276393

**Authors:** Scott Dryden-Peterson, Andy Kim, Arthur Y. Kim, Ellen C. Caniglia, Inga Lennes, Rajesh Patel, Lindsay Gainer, Lisa Dutton, Elizabeth Donahue, Rajesh T. Gandhi, Lindsey R. Baden, Ann E. Woolley

## Abstract

**Background:** In the EPIC-HR trial, nirmatrelvir plus ritonavir led to an 88% reduction in hospitalization or death among unvaccinated outpatients with early COVID-19. Clinical impact of nirmatrelvir plus ritonavir among vaccinated populations is uncertain.

**Objective:** To assess whether nirmatrelvir plus ritonavir reduces risk of hospitalization among outpatients with early COVID-19 in the setting of prevalent SARS-CoV-2 immunity and immune evasive SARS-CoV-2 lineages.

**Design:** Population-based cohort study analyzed to emulate a clinical trial utilizing two-stage, inverse-probability weighted models to account for anticipated bias in testing and treatment.

**Setting:** A large healthcare system providing care for 1.5 million patients in Massachusetts and New Hampshire during Omicron wave (January 1 to May 15, 2022) with staged access and capacity to prescribe nirmatrelvir plus ritonavir.

**Patients:** 30,322 non-hospitalized adults (87.2% vaccinated) aged 50 and older with COVID-19 and without contraindications to nirmatrelvir plus ritonavir.

**Measurement:** Primary outcome was hospitalization within 14 days of COVID-19 diagnosis.

**Results:** During the study period, 6036 (19.9%) patients were prescribed nirmatrelvir plus ritonavir and 24,286 (80.1%) patients were not. Patients prescribed nirmatrelvir were more likely to be older, have more comorbidities, and be unvaccinated. Hospitalization occurred in 40 (0.66%) and 232 (0.96%) patients prescribed and not prescribed nirmatrelvir plus ritonavir, respectively. The adjusted risk ratio was 0.55 (95% confidence interval 0.38 to 0.80, p = 0.002). Observed risk reduction was greater among unvaccinated patients and obese patients.

**Limitations:** Potential for residual confounding due to differential access and uptake of COVID-19 vaccines, diagnostics, and treatment.

**Conclusions:** The overall risk of hospitalization was already low (<1%) following an outpatient diagnosis of COVID-19, but this risk was 45% lower among patients prescribed nirmatrelvir plus ritonavir.

**Funding:** National Institutes of Health (P30 AI060354 and R01 CA236546).

## Introduction

The oral SARS-CoV-2 protease inhibitor nirmatrelvir, co-administered with the pharmacokinetic booster ritonavir, decreased the risk of progression to severe COVID-19 by 88% among unvaccinated high-risk patients with mild to moderate disease who enrolled when Delta (B.1.617.2) was the predominant circulating variant.^1^ Nirmatrelvir plus ritonavir was granted Emergency Use Authorization (EUA) in the United States in December 2021 for early COVID-19 and the national strategy encouraged broad utilization in increased risk individuals, regardless of vaccination status, to prevent hospital crowding.^2,3^ The World Health Organization recommended nirmatrelvir plus ritonavir in April 2022, but only for the highest risk individuals (>10% probability of hospitalization) and advised against use in most vaccinated and other lower risk individuals.^4^ Improved understanding of the clinical effectiveness of nirmatrelvir plus ritonavir is needed to inform individual and public health decisions, particularly among vaccinated individuals infected by Omicron (B.1.1.529, BA.1, BA.2, and other immune evasive sub-lineages).

## Methods

### Setting and Data

We utilized data from Mass General Brigham, a large non-profit, integrated healthcare system (1.5 million annual patients) that includes two large academic hospitals, seven community hospitals, and a network of ambulatory clinics and community health centers throughout Massachusetts and southern New Hampshire. During this period, Omicron lineages BA.1.1, BA.2, and BA.2.12.1 were predominant in the region.^5^ Nirmatrelvir plus ritonavir became available for prescription by Mass General Brigham providers for the highest risk individuals on January 21, 2022 and for all EUA-eligible patients on February 22, 2022. The study was approved by the Mass General Brigham Human Research Committee institutional review board and informed consent was waived.

Mass General Brigham hospitals and clinics utilize a shared electronic health record (EHR) (Epic Systems, Verona, WI). Dates of positive SARS-CoV-2 testing (including both polymerase chain reaction (PCR) tests as well as recorded home antigen tests that were documented in the EHR), inpatient admissions at one of the nine Mass General Brigham hospitals, patient demographics, comorbidities, concomitant home medications, COVID-19 treatments, COVID-19 vaccination status, and any deaths were obtained from an integrated COVID-19 data repository. Data was validated by manual review of medical records by two physicians. Similar to all other prescriptions, outpatient orders for nirmatrelvir plus ritonavir were required to be electronically transmitted to pharmacies per regulations in Massachusetts and New Hampshire (verbal and paper prescriptions were only permitted during system failures). Patients electronically prescribed courses of nirmatrelvir plus ritonavir were categorized as being in the treatment group.

### Patients

Individuals aged 50 years and older with a new diagnosis of COVID-19 (no prior positive molecular testing in preceding 90 days) between January 1 and May 15, 2022 and residing in Massachusetts or New Hampshire were included. Patient medical conditions, vaccination history, medications used at time of diagnosis, height and weight, self-reported race and ethnicity, and home zip code were obtained from the EHR. Recorded medical conditions and age were used to calculate the monoclonal antibody screening score (MASS), a comorbidity index that is predictive of COVID-19 hospitalization risk,^6,7^ for each patient. Patients were considered immunocompromised if they were receiving immunosuppressive medications (eg, prednisone, TNF inhibitors, calcineurin inhibitors, mTOR inhibitors, anti-CD20 antibodies), had an active malignancy, received a stem cell or solid organ transplant, or HIV infection irrespective of CD4 cell count.

Patients who received other approved treatments for early COVID-19— anti-SARS CoV-2 monoclonal antibodies (bebtelovimab or sotrovimab), molnupiravir, and outpatient remdesivir— were excluded. Patients with estimated glomerular filtration (eGFR) less than 30 mL/min or taking common medications where coadministration with nirmatrelvir plus ritonavir is not advised (i.e., cyclosporine, tacrolimus, everolimus, sirolimus, clopidogrel, rivaroxaban, amiodarone, carbamazepine, phenytoin, and ranolazine) were also excluded.

### Effectiveness

The primary objective was to assess the effectiveness of nirmatrelvir plus ritonavir in reducing the risk of hospitalization in the 14 days following an outpatient COVID-19 diagnosis among individuals 50 years and older (hospitalizations through May 29, 2022). While nirmatrelvir plus ritonavir became preferred therapy at Mass General Brigham in January 2022, limitations in supply, pharmacy locations, patient acceptance, provider awareness and clinical capacity to prescribe resulted in staged patient access. We compared patients prescribed versus patients not prescribed nirmatrelvir plus ritonavir to estimate the effectiveness of the treatment. After initial review of data, deaths occurring within 28 days of COVID-19 diagnosis were added as a secondary assessment of effectiveness.

The study included recorded COVID-19 infections occurring between January 1 and May 15, 2022 and any subsequent hospitalizations (through May 29, 2022) or deaths (through June 12, 2022).

### Statistical analysis

We sought to emulate a clinical trial utilizing observational data where nirmatrelvir plus ritonavir was assigned randomly among EUA-eligible individuals with early COVID-19 in the context of high prevalence of prior immunity and the current circulating variants. Date of COVID-19 diagnosis was used as start of period of risk in both study arms. In accordance with EUA requirements, patients receiving nirmatrelvir plus ritonavir started within first five days of symptoms. We anticipated bias related to vaccination, medical history, healthcare access, and other factors both at the stage of COVID-19 diagnosis (outpatient diagnosis versus diagnosis at time of admission) and at the stage of nirmatrelvir plus ritonavir prescription. To better achieve exchangeability between patients prescribed and not prescribed nirmatrelvir plus ritonavir, we utilized a two-stage, inverse-probability weighted design. Testing and treatment weights were separately generated using logistic models including *a priori* determined factors of age (50 to 64, 65 to 79, or 80 years and older), comorbidity score (MASS 1 to 3, 4 and 5, or 6 and higher), vaccination status (complete vaccination or incomplete/no vaccination), recency of last vaccine dose (less than 20 weeks or longer than 20 weeks per observed decreased protection against severe disease^8^), self-reported race and ethnicity (Hispanic or Latinx, White non-Hispanic or Latinx, Black non-Hispanic or Latinx, Asian non-Hispanic or Latinx, or Other/Unavailable), and socioeconomic vulnerability of residential address (75^th^ percentile and higher or lower than 75^th^ percentile of Massachusetts-specific Social Vulnerability Index^9^ averaged over zip code geography). The product of the testing and treatment weights were used to generate a pseudo-population^10^ to estimate the effect of nirmatrelvir plus ritonavir if all patients diagnosed with COVID-19 received their diagnosis as an outpatient and prescribing was not biased based on the included factors. A modified Poisson model using robust error variance^11^ and general estimating equations^12,13^ was used to estimate relative risk reduction with nirmatrelvir plus ritonavir compared with no treatment in the pseudo-population generated by the product of the testing and treatment weights.

## Results

### Study population and nirmatrelvir plus ritonavir prescription

Between January 1 and May 15, 2022, a total of 78,474 Mass General Brigham patients were diagnosed with COVID-19 of which 31,460 were 50 years and older and were eligible for this study. Among these patients, 1138 were first diagnosed at the time of hospital admission or death and were only included in the development of testing weights. The remaining 30,322 patients were diagnosed as outpatients and eligible for nirmatrelvir plus ritonavir (Figure 1).

**Figure 1.**
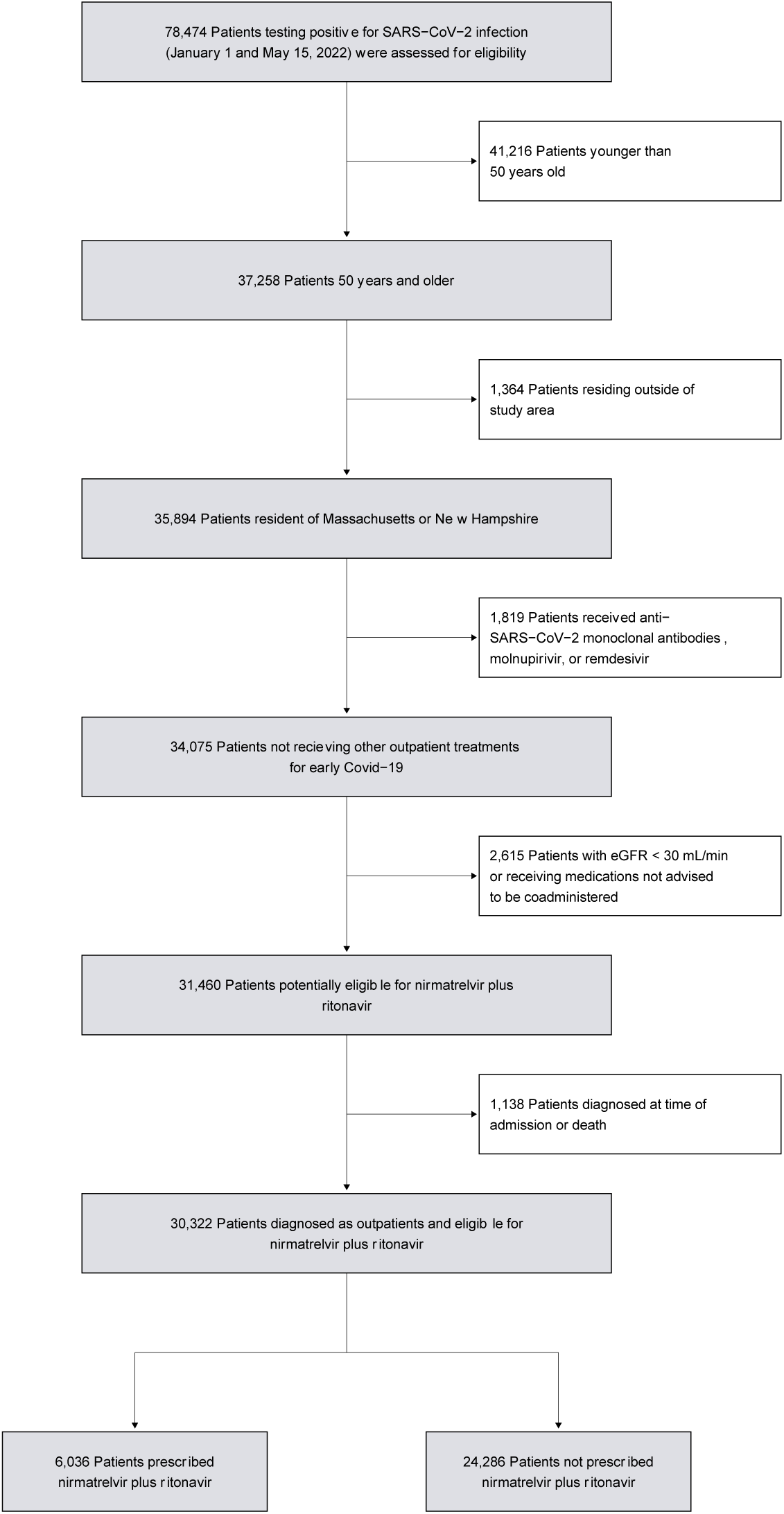
Study cohort.

The study period included an improving winter SARS-CoV-2 wave (January and February 2022) during which access to nirmatrelvir plus ritonavir was severely limited and an intensifying spring wave (April and May 2022) with greater access and provider willingness to prescribe nirmatrelvir plus ritonavir. Cases, hospitalizations, and prescriptions of nirmatrelvir plus ritonavir throughout the study period are shown in Figure 2.

**Figure 2.**
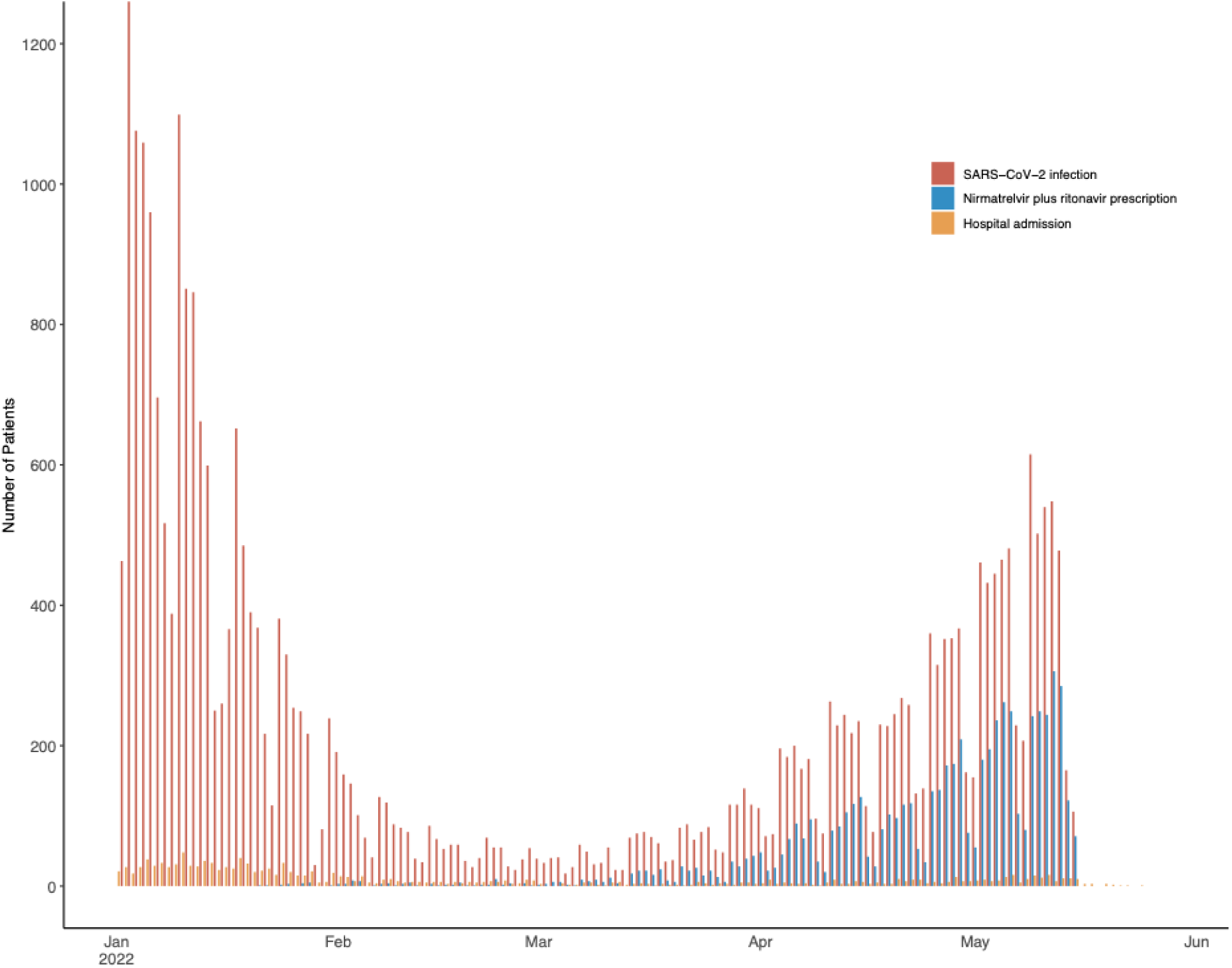
SARS-CoV-2 infection, treatment with nirmatrelvir plus ritonavir, and hospitalization among study patients. Infections and treatment initiation included from January 1 to May 15, 2022. Hospitalizations included through May 29, 2022.

The baseline characteristics of the analysis population are shown in Table 1. Nirmatrelvir plus ritonavir was prescribed to 6036 patients and was more likely to be prescribed to patients with older age, higher comorbidity scores, and completed vaccination. As evidenced by findings from the multivariable logistic model used to calculate treatment weights, patients residing in socioeconomically vulnerable zip codes (adjusted odds ratio [aOR] 0.68, 95% confidence interval [CI] 0.60 to 0.76, p<0.001) and patients reporting Hispanic or Latinx ethnicity (aOR 0.45, 95% CI 0.38 to 0.53, p<0.001), or Black race (aOR 0.43, 95% CI 0.34 to 0.52, p<0.001) were less likely to be prescribed nirmatrelvir plus ritonavir compared with higher income regions and patients reporting White race.

**Table 1.** Baseline characteristics of COVID-19 cases aged 50 and older (Jan 1 to May 15, 2022)

| Characteristic | Prescribed<br>nirmatrelvir<br>plus ritonavir | Not prescribed<br>nirmatrelvir plus<br>ritonavir | p |
| --- | --- | --- | --- |
| No. | 6036 | 24286 |  |
| Age (%) |  |  | <0.001 |
| 50 to 64 | 2745 (45.5) | 15084 (62.1) |  |
| 65 to 79 | 2693 (44.6) | 7391 (30.4) |  |
| 80 and older | 598 (9.9) | 1811 (7.5) |  |
| Male sex (%) | 2505 (41.5) | 9851 (40.6) | 0.189 |
| Race and ethnicity (%) |  |  | <0.001 |
| Asian | 171 (2.8) | 665 (2.7) |  |
| Black | 103 (1.7) | 1124 (4.6) |  |
| Hispanic or Latinx | 177 (2.9) | 1738 (7.2) |  |
| Other or unavailable | 206 (3.4) | 1416 (5.8) |  |
| White | 5379 (89.1) | 19343 (79.6) |  |
| High zip code SES vulnerability (%) | 422 (7.0) | 3020 (12.4) | <0.001 |
| Vaccination status (%) |  |  | <0.001 |
| Vaccinated and boosted | 4623 (76.6) | 11776 (48.5) |  |
| Vaccinated | 1048 (17.4) | 9002 (37.1) |  |
| Partially vaccinated | 96 (1.6) | 703 (2.9) |  |
| Unvaccinated | 269 (4.5) | 2805 (11.5) |  |
| Last vaccine dose more than 20 weeks prior (%) | 4821 (79.9) | 14990 (61.7) | <0.001 |
| Comorbidity score (%) |  |  | <0.001 |
| MASS 3 or less | 2821 (46.7) | 14940 (61.5) |  |

| Characteristic | Prescribed<br>nirmatrelvir<br>plus ritonavir | Not prescribed<br>nirmatrelvir plus<br>ritonavir | p |
| --- | --- | --- | --- |
| MASS 4 and 5 | 1089 (18.0) | 3611 (14.9) |  |
| MASS 6 or greater | 2126 (35.2) | 5735 (23.6) |  |
| Body mass index (BMI) (%) |  |  | <0.001 |
| BMI less than 25 or unavailable | 1851 (30.7) | 8880 (36.6) |  |
| BMI 25 to 30 | 2154 (35.7) | 7643 (31.5) |  |
| BMI 30 to 35 | 1250 (20.7) | 4753 (19.6) |  |
| BMI greater than 35 | 781 (12.9) | 3010 (12.4) |  |
| Immunocompromise (%) | 2836 (47.0) | 8749 (36.0) | <0.001 |
| Diabetes (%) | 1254 (20.8) | 4218 (17.4) | <0.001 |
| Heart disease or stroke (%) | 1020 (16.9) | 3054 (12.6) | <0.001 |
| Pulmonary disease (%) | 564 (9.3) | 1719 (7.1) | <0.001 |
| Bipolar, schizophrenia, and other disorders (%) | 128 (2.1) | 727 (3.0) | <0.001 |
| Depression and anxiety (%) | 1466 (24.3) | 4667 (19.2) | <0.001 |
| Hematologic malignancy (%) | 289 (4.8) | 694 (2.9) | <0.001 |
| Solid tumor malignancy (%) | 2224 (36.8) | 6538 (26.9) | <0.001 |
| Rheumatologic or inflammatory bowel disease (%) | 851 (14.1) | 2318 (9.5) | <0.001 |

Numbers are No. (%) unless otherwise noted. Immunocompromise includes patients with active malignancy and patients on immunosuppressive medications

### Hospitalizations and deaths

The primary endpoint of hospitalization within 14 days of incident COVID-19 infection occurred in 40 (0.66%) patients prescribed nirmatrelvir plus ritonavir and 232 (0.96%) who did not receive nirmatrelvir plus ritonavir. None of the hospitalizations among nirmatrelvir plus ritonavir recipients were attributable to the described rebound syndrome.^14^ The adjusted risk ratio was 0.55 (95% confidence interval [CI] 0.38 to 0.80, p = 0.002). The observed reduction in risk was similar across age, socioeconomic vulnerability, and comorbidities. However, nirmatrelvir plus ritonavir was associated with increased protective activity among patients with BMI 30 kg/m^2^ and higher compared with non-obese patients (p = 0.007). While difference did not reach statistical significance, nirmatrelvir plus ritonavir appeared to have greater activity among incompletely vaccinated individuals (p = 0.052), Figure 3.

**Figure 3.**
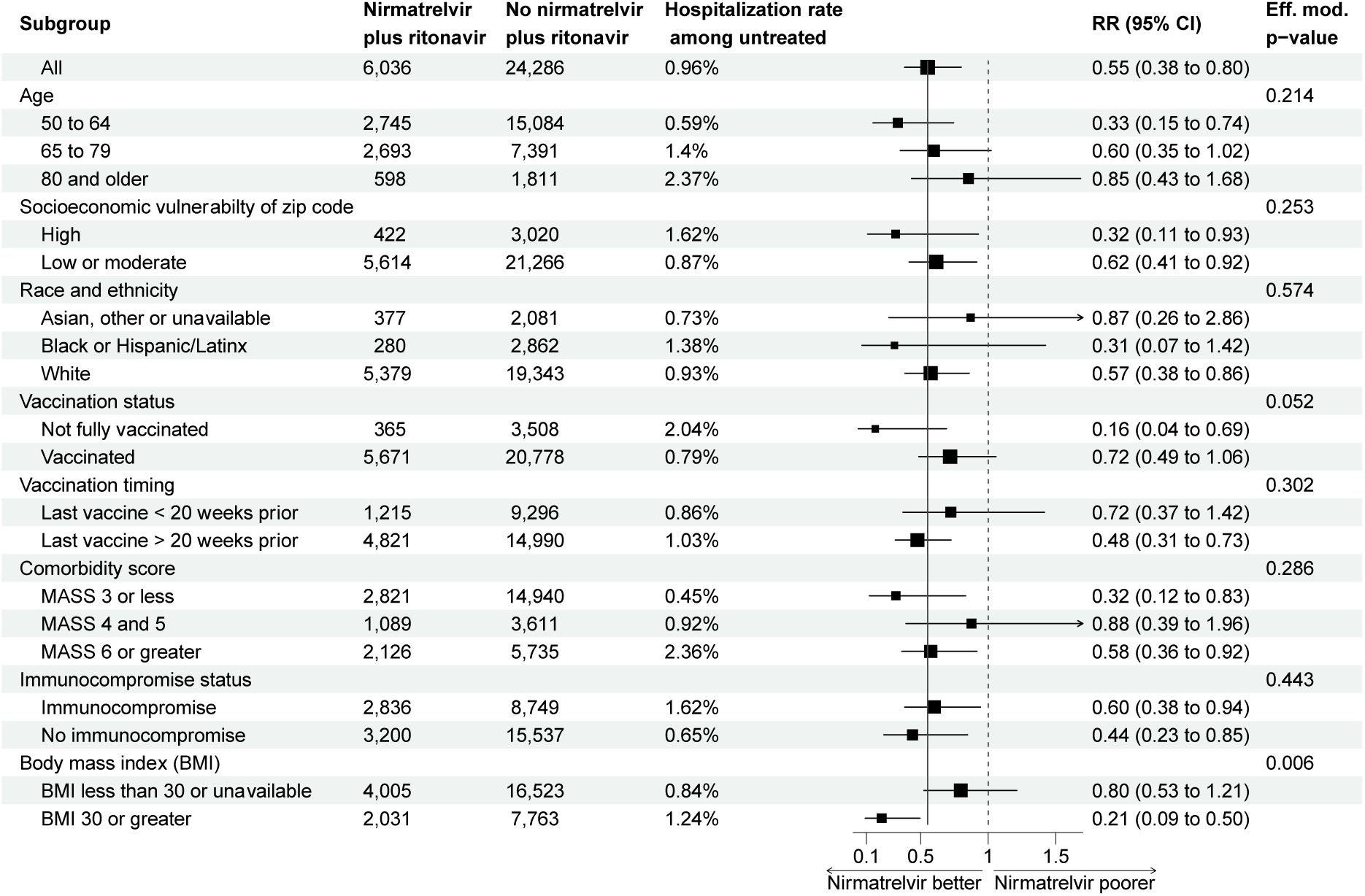
Subgroup analysis of the risk ratio of hospitalization comparing patients prescribed and not prescribed nirmatrelvir plus ritonavir. Estimate and confidence interval calculated from an inverse-probability weighted model performed within in strata. Effect modification p-value calculated from nested models.

A total of 39 deaths within 28 days of COVID-19 diagnosis were observed among the 30,322 study patients diagnosed as outpatients. All deaths occurred among patients who were not prescribed nirmatrelvir plus ritonavir. Thirteen of the patients died while hospitalized after COVID-19 diagnosis and 28 (74%) of the deaths occurred in vaccinated patients.

## Discussion

We emulated a clinical trial using observational data to evaluate the effectiveness of nirmatrelvir plus ritonavir among patients 50 years and older in preventing hospitalization in the context of high vaccination prevalence and an intense Omicron epidemic. The observed rate of hospitalization was low (less than 1%) among patients diagnosed with COVID-19 as outpatients, but we found that nirmatrelvir plus ritonavir was associated with a further 45% reduction in the risk of hospitalization.

In both the randomized trial and our analysis of observational data, there was a reduction in the rate of COVID-19 related hospitalization and death in the nirmatrelvir plus ritonavir arm. However, there are some important differences in study contexts that may account for a lower magnitude of risk reduction compared to the randomized controlled trial. The EPIC-HR study^1^ enrolled only unvaccinated individuals (median age 46) and had a 7% hospitalization rate in the placebo arm compared to our study which included mainly vaccinated patients (median age 62) and had an overall 1% hospitalization rate. It is possible that hospitalization risk cannot be reduced much further particularly among vulnerable patients. However, the 84% risk reduction seen in the subgroup analysis of unvaccinated individuals in our study was similar to the risk reduction observed in EPIC-HR of unvaccinated individuals. In addition, EPIC-HR excluded patients likely to require hospitalization within two days of randomization and less than 1% were admitted in this interval, however, 35% of the hospitalizations in the nirmatrelvir plus ritonavir arm of our study occurred within two days of prescription.

The results of our study also support the findings of two analyses of nirmatrelvir plus ritonavir in the same health system in Israel, though the methodology, inclusion criteria, and statistical analysis plans differ. These studies do not attempt to account for bias in early diagnosis or prescription, but rather balance the risk of hospitalization through standard Cox multivariable regression. Utilizing data from January and February of 2022, Najjar-Debbiny et al. similarly excluded patients diagnosed at time of admission and estimated a 46% reduction in hospitalization.^15^ In a more recent analysis, Arbel *et al*.^16^estimated a 77% reduction in hospitalization risk among patients 65 and older but greater risk reduction likely due to inclusion of patients diagnosed during admission and an immortal time bias.^16,17^

One of the strengths of our study is the large sample size which allowed us to estimate the risk reduction of nirmatrelvir plus ritonavir for specific subpopulations of interest. Overall, we observed a relatively consistent protective effect of nirmatrelvir plus ritonavir despite greatly varying hospitalization rates across groups. While unvaccinated patients appear to have experienced greater proportional reduction in risk (non-significant finding), vaccinated patients also had risk reduction. Similarly, there was a trend towards increased protection among those who received a vaccine dose more that 20 weeks prior. While not statistically significant the estimated risk reduction appeared to decrease with increasing age in contrast with results from study from Israel.^15^ Our use of a binary endpoint does not capture any effect of delaying admission among older patients at high risk that may have been detected in a time-to-event approach used by Najjar-Debbiny *et al*. Additionally, procedures in place in some skilled nursing facilities or elder home care programs encourage admission in context of COVID-19 pre-emptively or for infection control. While hospitalization risk is higher among obese individuals,^18^ the significant finding of enhanced risk reduction by nirmatrelvir plus ritonavir was unexpected and should be further explored.

The findings of this study should be interpreted in the context of several limitations. This was an observational retrospective study with residual confounding and selection bias. We attempted to minimize some of the selection bias by excluding patients whose initial diagnosis of COVID-19 coincided with their admission date as those patients were more likely to have had a delay in diagnosis and therefore more advanced disease at the time of diagnosis. This led to a lower hospitalization rate for the analyzed cohort than what was seen in the overall Mass General Brigham population during this time period. While the majority of hospitalizations for the study population would have been captured in the EHR given Mass General Brigham’s large healthcare system incorporating nine hospitals, hospitalizations outside our system were not included. Positive home antigen tests are incompletely captured in the EHR. While bias introduced was mitigated by two-stage design, residual bias was likely. Our analysis included an intention to treat approach but adherence to nirmatrelvir plus ritonavir may have been lower in our real-world study and some patients may not have even taken the prescribed treatment. potentially underestimating of the efficacy of this treatment.

The majority of COVID-19 admissions during the study period—1138 of 1410 hospitalizations— occurred among patients diagnosed at time of admission. Patients residing in socioeconomically vulnerable regions and patients identifying as Black or as Hispanic or Latinx were more than twice as likely to be diagnosed only during hospital admission. Similarly, among patients successfully diagnosed with COVID-19 as outpatients, these same groups experienced much lower rates of nirmatrelvir plus ritonavir prescription. To realize the public health potential of outpatient COVID-19 therapy we must address this gap and these disparities.

This study estimates a moderate effectiveness of nirmatrelvir plus ritonavir for preventing hospitalizations in a noncontrolled setting for a largely vaccinated population of individuals aged 50 or older. Furthermore, we demonstrate that the estimated benefit likely increases for individuals who are further out from their last vaccine dose, which is one of the key findings from this study with public health implications. Though its clinical impact may be reduced in vaccinated individuals, these results strengthen the expectation that nirmatrelvir plus ritonavir is an effective oral antiviral treatment in preventing individuals at increased risk for illness progression from requiring hospitalization due to COVID-19. It will be important to continue to assess the clinical efficacy of nirmatrelvir plus ritonavir in context of future variants and in relation to other therapeutic options.

## Data Availability

All data produced in the present study are available upon reasonable request to the authors.

## Acknowledgement

This work was made possible with help from the Harvard University Center for AIDS Research (CFAR), a funded program of the National Institutes of Health (P30 AI060354) and the National Cancer Institute (R01 CA236546). The contents of this manuscript are solely the responsibility of the authors and do not necessarily represent the official views of the National Institutes of Health or the institutions with which the authors are affiliated. The funding source had no role in the design and conduct of the study; collection, management, analysis, and interpretation of the data; preparation, review, or approval of the manuscript; and decision to submit the manuscript for publication. Drs. Dryden-Peterson and Woolley had full access to all the data in the study and take responsibility for the integrity of the data and the accuracy of the data analysis.

